# Mendelian Randomization Reveals the Role of HMGCR in Pulmonary Arterial Hypertension Treatment, Independent of LDL Cholesterol Concentration

**DOI:** 10.1101/2023.10.02.23295606

**Authors:** Shixun Wu, Jie Pu, Zhiguo Tang, Wei Zhang, Fangxia Hou, Na Wei, Qianwei Cui, Fuqiang Liu, Junkui Wang, Ying Lu, Tao Wei

## Abstract

**Introduction:** Specific lipid-reducing therapeutics, including statins, are known for mitigating cardiovascular diseases due to their comprehensive benefits including anti-inflammatory properties, antioxidative stress response, and enhancement of endothelial function. The objective of our study was to determine the causative impact of lipid-reducing agents (HMGCR inhibitors, PCSK9 inhibitors, and NPC1L1 inhibitor) on the outcomes of pulmonary hypertension via a two-sample Mendelian randomization (MR) analysis.

**Methods:** Two types of genetic tools were employed to estimate the exposure to lipid-lowering drugs, comprising expression quantitative trait loci of the drug’s target genes and genetic variations close to or within the target genes related to low-density lipoprotein (LDL cholesterol derived from a genome-wide association study). We utilized summary-data-based MR (SMR) and inverse-variance-weighted MR (IVWMR) methodologies for estimating effect sizes.

**Results:** SMR analysis indicated that elevated HMGCR expression correlates with increased pulmonary hypertension risk (β=-0.964, se=0.276). Yet, no evident causative link between HMGCR-regulated LDL cholesterol and COVID-19 hospitalization was observed in the IVW-MR analysis (β = -0.21, se= 0.17).

**Conclusions:** Our Mendelian randomization investigation unveiled a possible positive impact of lipid-lowering therapeutics on the prognosis of pulmonary hypertension. Importantly, no causal relation was established between LDL cholesterol and pulmonary hypertension.

## Introduction

Pulmonary hypertension, characterized by elevated pulmonary artery pressure, is a critical health issue leading to right ventricular collapse and subsequently, fatality.[1] Despite advancements in the understanding of the pathophysiological underpinnings of pulmonary hypertension over recent decades, the disease outlook remains bleak, marked by substantial mortality and morbidity rates.[2]

Abnormal lipid levels in the blood, a condition known as dyslipidemia, has been pinpointed as a crucial risk factor in the genesis of pulmonary hypertension.[3] The onset of dyslipidemia can precipitate alterations in pulmonary vascular structure and impede endothelial function, thereby intensifying the severity of pulmonary hypertension.[4] Given these factors, it is plausible to posit that reducing lipid levels could enhance the prognosis of pulmonary hypertension.[2][3] Specifically targeted lipid-reducing therapeutics, including statins, have been found to ameliorate the outlook of cardiovascular diseases, potentially via their anti-inflammatory, antioxidative stress, and endothelial function-improving properties.[5]

Nonetheless, the exact contribution of targeted lipid-reducing agents to the prognosis of pulmonary hypertension is yet to be completely elucidated.[6] While conventional observational research has suggested potential beneficial effects of these agents, the results require additional verification owing to possible influences of confounding elements and reverse causality.

Mendelian randomization, a genetics-driven methodology, can aid in discerning potential causative associations.[7] This technique employs genetic variance as a pivotal variable, thereby evading confounding influences and reverse causality.[7] As such, we opted for a Mendelian randomization approach to explore the linkage between targeted lipid-lowering medications and the prognosis of pulmonary hypertension.

The objective of our research was to ascertain whether targeted lipid-lowering therapeutics could enhance the prognosis of pulmonary hypertension through a Mendelian randomization methodology. We hypothesize that the utilization of lipid-reducing medications might correlate with an improved prognosis of pulmonary hypertension. If validated, this finding could contribute significantly to the management and therapy of pulmonary hypertension.

## Materials and methods

### Study design

This dual-sample Mendelian randomization (MR) investigation utilized publicly accessible aggregate data from Genome-Wide Association Studies (GWAS) and Expression Quantitative Trait Loci (eQTLs) studies (Supplementary file 1—Table 1). Each of these studies received approval from pertinent institutional review boards, and all participants gave their informed consent.

### Selection of genetic instruments

This research included three categories of FDA-sanctioned lipid-lowering pharmaceuticals as exposures: inhibitors of HMGCR, PCSK9, and NPC1L1. As indicated in Table 1, we utilized accessible eQTLs for the target genes of these drugs (i.e., HMGCR, PCSK9, and NPC1L1) as indicators of exposure to each lipid-lowering medication. The summary data of eQTLs was sourced from the eQTLGen Consortium (https://www.eqtlgen.org/) or the GTEx Consortium V8 (https://gtexportal.org/), the specifics of which are elaborated in Supplementary file 1— Table 1. We pinpointed prevalent (minor allele frequency [MAF] >1%) eQTLs single-nucleotide polymorphisms (SNPs) significantly (p < 5.0 × 10-8) linked with the expression of HMGCR or PCSK9 in blood, and the expression of NPC1L1 in subcutaneous adipose tissue, given the absence of eQTLs in blood or other tissues for NPC1L1 at an equivalent significance level. The formation of genetic instruments in this study was based solely on cis-eQTLs, defined as eQTLs situated within 1 Mb on either side of the encoded gene. In a secondary approach to validate the observed relationship using eQTLs as an instrument, we suggested an additional instrument by choosing SNPs within a 100 kb range from each drug’s target gene, which were associated with LDL cholesterol levels at a genome-wide significance level (p < 5.0 × 10-8), as surrogates for the exposure to lipid-lowering medications. We identified these SNPs using GWAS summary data for LDL cholesterol levels from the Global Lipids Genetics Consortium (GLGC), with a sample size of 173,082, including only common SNPs (MAF >1%) (Supplementary file 1 Table 1).[8] We focused on single-nucleotide polymorphisms (SNPs) that were significantly linked with LDL cholesterol levels (p < 5.0 × 10-8) from genome-wide association studies (GWAS). These SNPs served as instrumental variables to assess the causality of LDL cholesterol levels (p < 5 × 10-8, r 2 < 0.001 and clump distance >10 000 kb). For the SNPs that were absent in the summary-level GWAS data for pulmonary hypertension, we resorted to linkage disequilibrium (LD) proxy SNPs (r^2 > 0.80). Moreover, palindromes having moderate allele frequencies were excluded from the study. Among the crucial SNPs, any strongly tied with potential confounding factors of pulmonary hypertension - such as cardiovascular disease, drug use, metabolic disorders, and endocrine disorders - were also eliminated, leaving only 45qualified SNPs. To identify any potential bias in the weak instrumental variable, we computed the F-statistic using the formula: F = R2(n - k - 1)/k(1 - R2), where R2 stands for the variance proportion of exposure elucidated by chosen genetic instruments, n is the sample size of exposure GWAS, and k represents the quantity of selected genetic instruments. An average F-statistic > 10 indicates the appropriateness of the instrumental variables used.

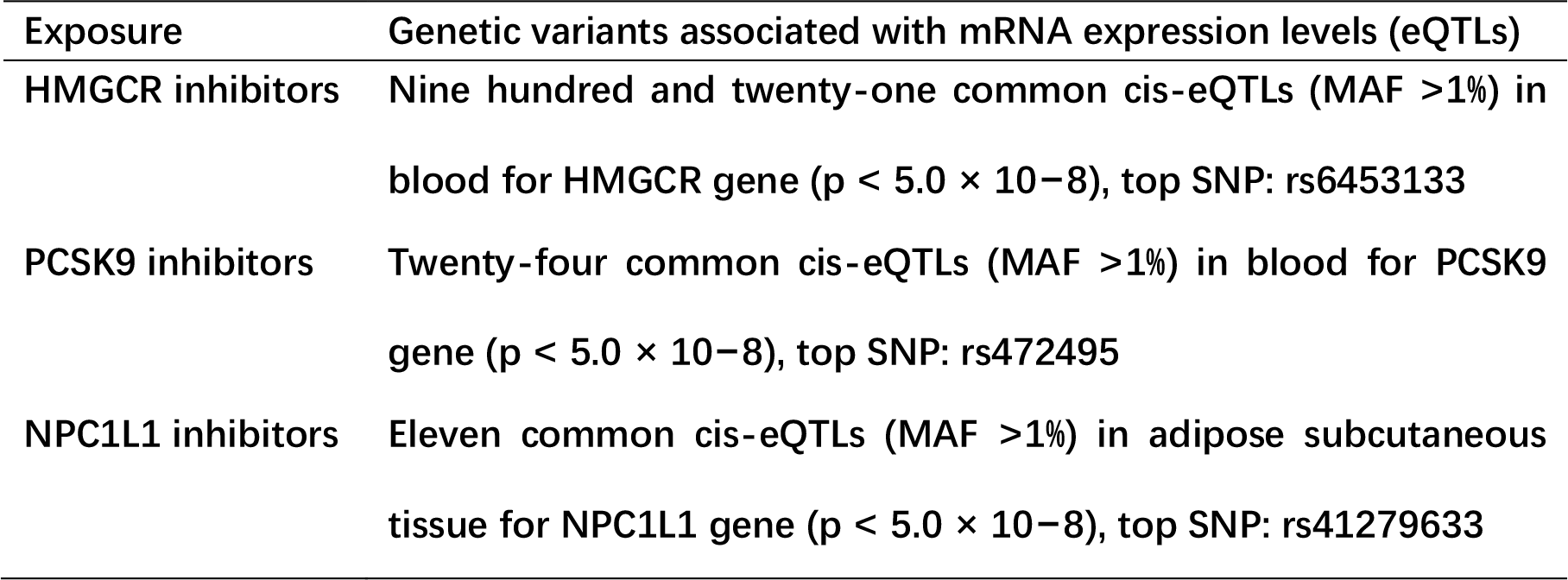

### Outcome sources

We sourced our data from the Gwas Catalog (https://www.ebi.ac.uk/gwas/home/), a resource built by the European Molecular Biology Laboratory. The summary-level data included in this research under the GWAS ID: GCST007228, Details pulmonary arterial hypertension with 2085 cases and 9659 non-cases.[9]

### Statistical analyses

#### MR analysis

We employed the summary-data-based MR (SMR) methodology to deduce effect estimates when leveraging eQTLs as a tool. This approach explores the relationship between gene expression levels and the targeted outcome by harnessing summary-level information from GWAS and eQTL research.[10] The harmonization and examination of alleles were conducted using SMR software, version 1.03, accessible at https://cnsgenomics.com/software/smr/#Overview. To amalgamate effect estimates when utilizing genetic variants linked with LDL cholesterol levels as a tool, we implemented the inverse-variance-weighted MR (IVW-MR) approach. The alignment and study of alleles were accomplished via the TwoSampleMR package within the R software platform, version 4.2.0.

#### Sensitivity analysis

We evaluated the potency of SNPs employed as instrumental variables via the F-statistic, opting for SNPs with an F-statistic over 10 to curb the influence of weak instrument bias.[11] To validate both genetic instruments, we conducted positive control analyses. Given the established impact of lipid-lowering drugs on LDL cholesterol reduction, we scrutinized the relationship between the exposure variables and LDL cholesterol levels as a positive control study for the eQTLs instrument.

Using the SMR approach, we employed the Heterogeneity in Dependent Instruments (HEIDI) test within the SMR software to assess whether the noted association between gene expression and outcome stemmed from a linkage scenario.[10] The HEIDI test with a p-value less than 0.01 suggests the association is likely attributable to linkage.[12] A single SNP could influence the expression of multiple genes, potentially leading to the occurrence of horizontal pleiotropy. To evaluate the risk of horizontal pleiotropy, we pinpointed additional neighboring genes (within a 1 Mb window) whose expression was significantly tied to the genetic instrumental variant. We then executed an SMR analysis to investigate if the expression of these genes correlated with pulmonary hypertension.

In the IVW-MR approach, we employed the Cochran Q test to check for heterogeneity, with a p-value less than 0.05 signifying evidence of heterogeneity.[13] We employed MR-Egger regression and Mendelian Randomization Pleiotropy RESidual Sum and Outlier (MR-PRESSO) analysis to evaluate the potential horizontal pleiotropy of the SNPs utilized as instrumental variants. Within MR-Egger regression, the intercept term serves as a handy indicator of directional horizontal pleiotropy, with a p-value less than 0.05 demonstrating evidence of horizontal pleiotropy.[11] MR-PRESSO analysis is capable of recognizing horizontal pleiotropic outliers and offering adjusted estimates, with a p-value less than 0.05 for the Global test denoting the existence of such outliers.[14]

## Results

### Genetic instruments selection and pulmonary arterial hypertension

From the eQTLGen or GTEx Consortium, we recognized 921, 24, and 11 cis-eQTLs for the drug target genes HMGCR, PCSK9, and NPC1L1, correspondingly. For each drug’s target gene, we chose the most impactful cis-eQTL SNP as a genetic instrument (Refer Table 1, Supplementary file 1—Table 2). The F-Statistics for all the instrument variants exceeded 30, indicating a probable mitigation of weak instrument bias in our investigation (Supplementary file 1-Tables 2). Leveraging the data from pulmonary hypertension GWAS, we used a collective count of 2085 cases and 9659 controls to probe the correlation with pulmonary hypertension.

### Primary analysis

From the SMR analysis, HEIDI testing proposed that all noticeable associations were unrelated to a linkage (p > 0.01), with an exception of the link between HMGCR expression and pulmonary arterial hypertension(p = 0.00048).(Table3) By analyzing 45 SNPs correlated with LDL cholesterol levels, we could not locate any significant evidence pointing to a potential causal impact of LDL cholesterol levels on pulmonary hypertension susceptibility (β=-0.21, se=0.17, p=0.21). Concurrently, MR-Egger regression (β = -0.23, se = 0.44, p = 0.69) and a weighted median technique (β=-0.32, se=0.26, p=0.21) resulted in comparable risk approximations, though the connection wasn’t statistically meaningful. Moreover, no heterogeneity was evident as the Cochran Q test produced a p-value of 0.89 for MR-Egger and a p-value of 0.91 for IVW.(Supplementary file 1-Figure)

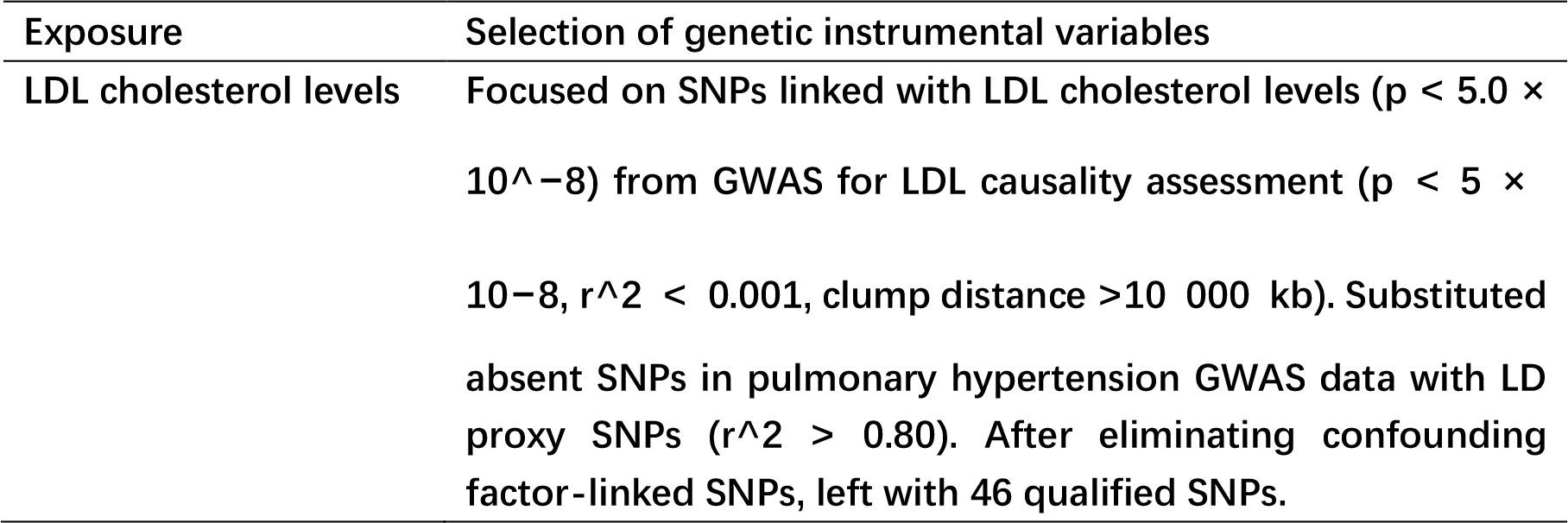

**Table 3.** SMR association between expression of gene HMGCR, PCSK9, or NPC1L1 and COVID-19 outcomes.

| GENE | SMR association |  |  |
| --- | --- | --- | --- |
|  | beta | se | p-value |
| HMGCR | -0.964 | 0.276 | 0.00048 |
| PCSK9 | -0.141 | 0.192 | 0.462 |
| NPC1L1 | 0.036 | 0.102 | 0.723 |

### Results from two sample MR

**Figure 1.**
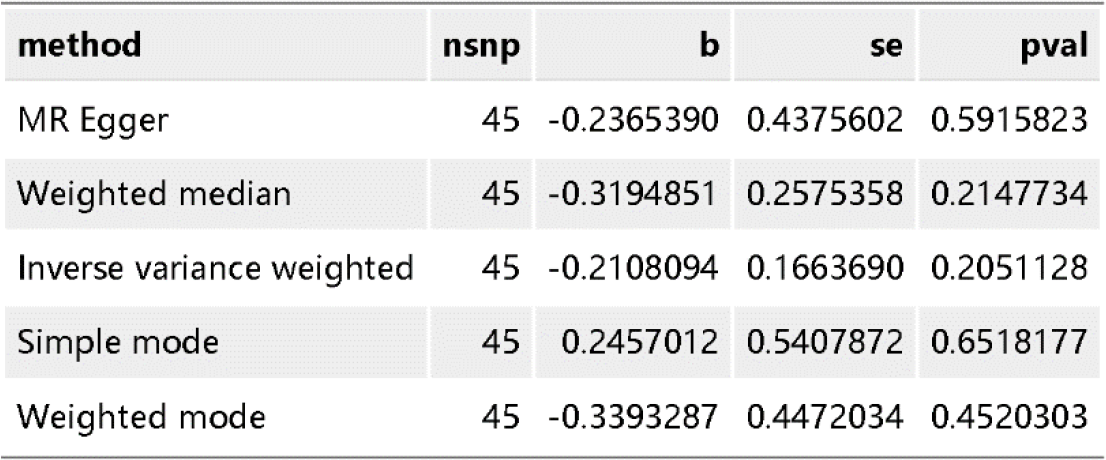
causal impact of LDL cholesterol levels on pulmonary hypertension.

## Conclusions

To conclude, our Mendelian randomization investigation identified a probable advantageous effect of lipid-lowering medications on the enhancement of pulmonary hypertension prognosis. Importantly, there was no causal linkage observed between LDL cholesterol and pulmonary hypertension. The pioneering employment of genetic instruments effectively reduced confounding and reverse causality, thus providing dependable evidence. Such discoveries could hold significant clinical implications, potentially instigating a paradigm shift in the therapeutic approaches for pulmonary hypertension. Nonetheless, in spite of the promising results, we underline the necessity for additional corroborating studies to solidify these findings. This research underscores the potency of genetic epidemiology in unraveling intricate disease mechanisms and therapeutic interventions, paving the path towards personalized medicine for treating pulmonary hypertension.

## Discussion

Our investigation suggests potential efficacy of HMGCR, a targeted point of action for lipid-lowering medications, in the treatment of pulmonary hypertension. Yet, no causal association was observed between LDL cholesterol levels and pulmonary hypertension when LDL cholesterol was used as the exposure variable. This could imply that the functionality of HMGCR isn’t confined to the reduction of LDL cholesterol levels. Other biological pathways, such as anti-inflammatory responses or impacts on vascular endothelial function, could also be in play.[15]

Nevertheless, our study possesses certain limitations. Initially, our sample size may not have been adequate to detect the mild association between LDL cholesterol levels and pulmonary hypertension. Secondly, potential biases might exist in our study design, for instance, possibly insufficient control of confounding variables. Finally, biases could be inherent in our data sources, such as measurement inaccuracies or reporting bias. These restrictions might have influenced our results; hence our findings necessitate validation through larger samples.

Nonetheless, our study indicates crucial paths for upcoming research. Firstly, it is crucial to delve deeper into the functioning of HMGCR to comprehend its influence on the onset and progression of pulmonary hypertension. Secondly, contemplating Mendelian randomization studies using other exposure variables, like expression levels of HMGCR or functional variants, could be worthwhile.

From a clinical perspective, if HMGCR indeed exhibits a therapeutic effect on pulmonary arterial hypertension, lipid-lowering drugs could emerge as a novel therapeutic approach. Yet, this proposition necessitates validation via clinical trials.

Lastly, certain discrepancies exist between our results and those from other studies. For instance, some investigations have revealed a positive correlation between LDL cholesterol levels and the risk of pulmonary arterial hypertension.[16] This could be attributed to the fact that these studies employed an observational design, while ours utilized a Mendelian randomization design, which have contrasting assumptions and methodologies. Hence, other studies need to corroborate our findings.

### Study strengths

The current investigation boasts several advantages. Of particular note is the employment of a Mendelian randomization approach via genetic instruments to replace lipid-lowering drug exposures, hence mitigating confounding bias and circumventing reverse causality. Significantly, we incorporated two distinct categories of genetic instruments to substitute the drugs being investigated, thereby facilitating the cross-validation of impact approximations. Furthermore, we carried out comprehensive sensitivity analyses to verify the accuracy of the genetic instruments and the suppositions underpinning the Mendelian randomization research. The considerable sample size of our research bolsters the statistical potency and trustworthiness of our conclusions. Our results hold the potential to furnish invaluable insights regarding the role of lipid-lowering medications in PH outcomes, which may inform subsequent therapeutic strategies for this debilitating ailment. We discovered that while targeted lipid-lowering drugs potentially possess the capability to treat pulmonary arteries, two-sample Mendelian randomization indicated that LDL cholesterol doesn’t incite pulmonary hypertension, underscoring the potential significance of our research.

### Study limitations

The current research has a number of shortcomings. Primarily, the absence of eQTLs for NPC1L1 in blood impeded our examination of the correlation between NPC1L1 expression in blood and pulmonary hypertension outcomes. Furthermore, the non-existence of eQTLs for these target genes in the liver (a principal tissue linked to lipid metabolism) might have strengthened the evidence for the observed associations. The eQTL investigations of PCSK9 and NPC1L1 in GTEx utilized comparatively small sample sizes, potentially affecting the statistical robustness of the results pertaining to PCSK9 or NPC1L1 inhibition. Secondly, the usage of pooled-level data prevented us from conducting subgroup analyses. Hence, additional MR studies employing individual-level data are required to yield more granular information. Thirdly, despite conducting diverse sensitivity analyses to verify the MR study hypotheses, the complete elimination of confounding bias and/or horizontal pleiotropy remained elusive. Fourthly, we advocate for an increase in randomized clinical trials and fundamental experiments to elucidate more detailed relationships between targeted lipid-lowering medications and LDL cholesterol levels in pulmonary hypertension. Lastly, the eQTLs and GWAS data utilized in this study primarily originated from populations of European ancestry. As such, these findings must be cautiously interpreted when extended to other populations.

## Data Availability

The data used in our study is sourced entirely from the Opengwas website, which is a public, ethically-reviewed database.

https://gwas.mrcieu.ac.uk/

